# Remote assessment of functional mobility within a community telehealth falls prevention program: interrater reliability and rating agreement

**DOI:** 10.64898/2026.01.10.25342852

**Authors:** N. Dionne, M Bilodeau, J O’Neil

## Abstract

In Canada, francophone older adults have difficulty accessing healthcare, which includes falls prevention. Technology may be used to improve access. We examined the interrater reliability of conducting the Timed Up and Go (TUG) and Five Times Sit to Stand (FTSTS) remotely compared to in-person within the community. Fifty-seven participants completed assessments with one in-person and one remote assessor simultaneously, while six participants completed the procedure with two remote assessors. Inter-rater reliability was evaluated between simultaneous assessment modes, and four participants completed single-rater assessments. Our study suggests an agreement between remote, or in-person assessments for the identification of older adults at increased risk of falls. These preliminary findings show remote assessments could be a good alternative to in-person assessments and can be done safely using technology for French-speaking older adults, potentially improving access to fall prevention services for older adults who face mobility, geographic or linguistic barriers.

**Plain language title:** *Comparing remote walking and balance assessments with in-person assessments.:* In Canada, francophone older adults have difficulty accessing healthcare, which includes falls prevention. Technology may be used to improve access to such services. However, comparing walking and balance assessments in-person versus remotely has not been fully studied in francophone older adult communities. We therefore wanted to study this comparison. Two clinical tests were completed remotely by physiotherapists and in-person by individuals with varied healthcare backgrounds. The differences in scores between an assessor in-person and one using videoconference were recorded. This helped document if results from an in-person assessment are the same as the ones done remotely. Sixty-seven older adults completed assessments either in-person or remotely. Our study suggests an agreement between simultaneous remote and in-person assessment scoring, correctly identifying the same participants at risk of falls. These preliminary findings show remote assessments could be a good alternative to in-person assessments and can be done safely using technology for French-speaking older adults, potentially improving access to fall prevention services for older adults who face mobility, geographic or linguistic barriers.

## Introduction

Primary falls prevention programs tailored for older adults have the potential to reduce the risk of falls, prevent secondary injuries such as fractures and head injuries, prevent loss of daily function, and improve social participation (Montero-Odasso et al., 2022; Pillay et al., 2024), thereby contributing to decreasing the overall burden on the healthcare system. According to World Guidelines for Fall Prevention, most programs should be multifactorial, including an educational component and an exercise program that is intensive enough to spark change (Montero-Odasso et al., 2022). Montero-Odasso et al. (2022) also recommend including a rigorous falls risk screening as part of prevention programs. In Canada, current falls prevention programs have been shown to be effective in improving mobility, balance, and knowledge around falls risk factors (Fauchard & Le Cren, 2009; Savard et al., 2018; Scott et al., 2011; Taing & McKay, 2017). Beyond screening and multifactorial interventions, it is essential to consider the needs of the community, the environmental context, and the available infrastructure for successful implementation (O’Neil et al., 2024). For example, falls prevention programs, such as “Marche vers le Futur” (MVF)(O’Neil et al., 2024; Savard et al., 2018) are effective primary prevention programs targeting older adults living in a minority situation.

In Canada, Francophones living in a minority situation are defined as French-speaking individuals living in provinces and territories outside of Quebec (De Moissac & Bowen, 2019). Considering the impact of linguistic barriers on quality of care (De Moissac & Bowen, 2019; Drolet et al., 2015), training local community program facilitators to deliver linguistically appropriate services that fulfill community needs is crucial and contributes to healthcare system efficiency (Savard et al., 2018). It has been shown feasible to deliver such programs in-person, remotely, synchronously, asynchronously, in-hospital settings, private clinics, community centres, and at home (Jacobson et al., 2021; Moriichi et al., 2022; O’Neil et al., 2024). However, these studies focused on remote intervention delivery rather than remote assessments completed within francophone minority communities. While some programs have included the completion of in-person pre- and post-program assessments to either determine eligibility or document changes in fall risks, this may require additional human or financial resources and can limit access for people with limited transportation or who live far from main centres (Belanger et al., 2023). Exploring digital health methods to assess program eligibility or pre and post program changes from a community-based perspective is necessary. Functional mobility assessments can be completed by any trained healthcare professional, however it may be difficult to access such a service in one’s language of choice, depending on geographical location. For certain health conditions, remote assessments have been shown to have similar validity as when they are conducted in-person (Lee et al., 2024). As such, remote functional mobility assessments could be a good option to improve access to an initial screening (Mohammed et al., 2021; Patterson et al., 2022), positively impact triage efficiency and allow people to be assessed in their living environments, as opposed to unfamiliar locations such as a healthcare facility (Pelicioni et al., 2022).

However, Internet connectivity, limited digital literacy, and limited information around the reliability and validity of assessments conducted remotely, when compared to in-person, are often reported barriers to the use of remote assessments (Farzad et al., 2023; Franco et al., 2024). A common misconception amongst healthcare professionals is that older adults may present with digital literacy limitations, preventing them from adequately using technology. This creates a digital health divide that needs to be addressed so that older adults are not disadvantaged in accessing remote options (Mace et al., 2022). “Marche vers le Futur” (MVF), a community telehealth falls prevention program designed to be delivered remotely to French-speaking older adults by trained local community health professionals, has demonstrated effectiveness in reducing fall risk factors (Savard et al., 2018) and showcased important gains in capacity building around the use of telehealth across Canada (O’Neil et al., 2024). Informed by the results of a previous study (O’Neil et al., 2024), trained program facilitators wanted to determine if the use of remote assessments provided a reliable and valid fall-risk screening alternative for people living in remote and rural areas. While remote assessments were already being conducted by MVF facilitators to determine program eligibility, demonstrating feasibility, their reliability and validity had not been objectively examined compared to in-person assessments with individuals from francophone minority communities.

### Reliability and validity

Recent studies have found good to excellent levels of reliability and validity between remote and in-person fall risk assessment tests, such as the Sit-to-Stand, Timed-Up-and-Go and Berg Balance Scale (Jasper et al., 2025). However, the geographical and linguistic context of Francophones living in a minority situation is not usually taken into consideration (González-Calvo et al., 1997; International Test Commission (ITC), 2019). Indeed, assuming cultural universality and not considering potential language barriers can result in unreliable assessment results (González-Calvo et al., 1997). Beyond linguistic differences, other aspects should be considered to ensure feasibility and reliability of remote assessments including access to reliable high-speed Internet, availability of appropriate technology at home (e.g., a computer or tablet), as well as digital literacy. Such factors can vary greatly across Canada’s diverse demographic and geographic contexts, which include many francophone communities located in provinces where English is the majority language. While validity and reliability studies of remote balance assessments in older adults exist, to our knowledge, none of these studies have explored the inter-rater reliability and rating agreements in the context of a falls prevention program for francophone minority communities living in a minority situation or rural, remote areas.

## Objectives

The objectives of this study were to examine: (1) the inter-rater reliability of remotely administered TUG and FTSTS assessments, (2) the inter-rater reliability of remote versus in-person administration, and (3) the concurrent agreement between the ratings of remote fall-risk screening compared with in-person screening among French-speaking older adults from francophone communities across Canada, including francophone minority communities.

## Methods

We used a descriptive pre-post multiple rater design to document inter-rater reliability as well as examine agreement between simultaneous ratings of 1) remote mobility assessments when compared with in-person assessments conducted simultaneously, and 2) remote mobility assessments when compared with remote assessments conducted simultaneously. All assessment sessions throughout this study were held in French. This study was part of a larger study comparing remote program delivery from a health equity perspective which included francophone minority communities across Canada (University of Ottawa board of ethics # H-07-23-9360 and Bruyère Health Research Ethics Board M16-23-056).

### Participants

Between January 2023 and June 2025, we recruited participants aged 55 years and up from different provinces who were interested in completing a telehealth falls prevention program in-person at a community centre or remotely, from their homes. Mindful of diverse settings across francophone communities, we recruited participants through various organizations including local francophone community health centres, and health organizations providing services to francophone communities. The lower age threshold of 55 years was used for this study because it corresponds to the eligibility criteria for the MVF program. To be included in this study, participants were screened via telephone for the following inclusion criteria: to speak French fluently, be interested in falls prevention, and be able to walk 3 meters without a gait aid or assistance. Participants who met these criteria provided written informed consent before data collection. A target sample size of 56 participants was estimated by accounting for a 15% attrition rate and based on a desired 0.8 power with an alpha of 0.05, and an inter-rater expected agreement (Kappa) of 0.9 (https://wnarifin.github.io/ssc/sskappa.html).

One of two trained physiotherapists who completed the MVF program facilitator training collected functional mobility data for each participant. The pre- and post-program shared assessment procedures included the Timed Up and Go (TUG) (Christopher et al., 2021), and the Five Times Sit to Stand (FTSTS) (Bohannon, 2006). The TUG is a reliable measure of functional mobility, where the person is asked to stand from a standard chair with a seat height of approximately 46 cm and armrest height of approximately 64 cm, walk 3 meters at a comfortable speed, turn around, return to the original chair, and sit (Christopher et al., 2021). This clinical assessment is recommended by the World Falls Prevention Guidelines as an objective measure to assess the risk of falls (Montero-Odasso et al., 2022). While a gait aid is allowed for this test, one of the inclusion criteria for the study was that participants safely walk without a gait aid on a 3m distance. Participants who completed the assessment therefore did not use a gait aid. The FTSTS is a functional strength test measuring the ability to get up from a chair and sit back down, with arms crossed across the chest, as fast as possible five times(Bohannon et al., 2015; Buatois et al., 2008). Normative values for older adults are: 11.4 s for the 60-69 age group; 12.6 s for the 70-79 years age group and 14.8 s for the 80-89 age group (Bohannon et al., 2015).The trained in-person program facilitators had backgrounds in physiotherapy, nursing, health promotion education, social work, kinesiology and medical diagnostic imaging. Each in-person assessment was completed at a local community centre.

To begin each assessment, participants received the verbal instruction "un, deux, trois, Go." The assessors started their timer at the "Go" instruction. For the TUG, the timer was to be stopped when the participant had returned to the initial sitting position whereas for the FTSTS, the timer was to be stopped once the participant was in a standing position after the fifth sit to stand repetition. No Internet connection delays required the remote assessors to start the timer at a different time than the verbal "Go" instruction given to the participants. Assessors were to start the timer as soon as they started to see the participant move if there were to be a connection delay.

To ensure safety for the 10 home remote-only assessments, standardization procedures and validity of assessment, participants were asked to measure the distance required for the TUG while being on the videoconference call with the assessors. Prior to the assessment, participants were advised to wear proper footwear, have another person physically present with them for the assessment, and emergency contact information as well as the participants’ address had been documented by the assessors. The participants’ phone numbers were also documented in anticipation of potential loss of Internet connection. Even though participants were given the "un, deux, trois, go" instruction to begin each test, remote assessors were to start their timers when they first saw the participants start moving as opposed to using the verbal "Go" instruction if there was a connection delay.

### Data collection

We collected participants’ socio-demographic data, which included: age, sex at birth, gender identity, native language, preferred spoken language, and origin. We screened for adverse events and technological or physical difficulties encountered during test administration. Adverse events included any incident compromising participants’ safety during or immediately after testing (e.g., falls, loss of balance, dizziness, pain, or inability to safely complete the assessment). Difficulties included physical limitations, difficulty following instructions and technological issues affecting remote assessments.

Two assessment conditions were selected for the inter-rater reliability and concurrent agreement comparisons: 1) in-person assessments compared with simultaneous remote assessments, and 2) remote assessments compared with simultaneous remote assessments. These conditions were chosen to reflect the different contexts in which falls prevention programs such as MVF are delivered. While some participants can access in-person assessments at a local community centre, others may rely entirely on remote assessments due to geographical location, mobility restrictions or limited availability to trained, French-speaking healthcare professionals. Since MVF is delivered both in-person and remotely, establishing the reliability of both assessment approaches was essential.

### Simultaneous remote assessment agreement protocol between two remote assessors

Two assessors were remotely evaluating each participant simultaneously. The simultaneous remote assessment option was provided to participants who completed the home version of the program when an in-person assessment was not feasible. Remote assessors were consistently French-speaking trained physiotherapists and program facilitator trainers with years of experience in remote care. For this mode of assessment, participants were instructed to use a device equipped with a camera such as a desktop, laptop or tablet. The camera was to be placed in such a way that the remote assessor could see the participant from head to toe throughout the required tasks. Participants were also instructed to minimize background noise to facilitate communication with the assessors and avoid harsh backlighting as much as possible. Sessions were all held on Zoom or Teams video platforms, depending on available technological infrastructure in each community. Participants were instructed to use a standard chair with a seat height of approximately 46 cm and armrest height of approximately 64 cm. When two remote assessors were available, they conducted the pre- and post-program assessment procedures simultaneously during the same session. To ensure safety, standardization procedure, and validity of assessment; participants were asked to measure the distance while being on the videoconference call with the assessors. Each test was performed twice, however only the second attempt of the TUG was recorded while both FTSTS tests were documented.

### Simultaneous remote and in-person assessments procedures

For this mode of assessment, one assessor was remote while the other was in-person with the participant. The remote assessors were the same French-speaking physiotherapists described in the previous section whereas the in-person assessors were French-speaking trained program facilitators with various healthcare professional backgrounds. The technology requirements were the same as the ones for the remote only assessments as one of the assessors still had to evaluate the participants from a separate location.

We examined the inter-rater reliability of assessment during a shared bimodal administration: 1) two trained physiotherapists who completed the MVF program facilitator training were responsible for collecting pre- and post-program assessment data remotely, and 2) trained program facilitators with backgrounds in physiotherapy, nursing, health promotion education, social work, kinesiology or medical diagnostic imaging were responsible for collecting pre- and post-program assessment data, in-person. Each in-person assessment was completed at a local community centre. This means that for the 57 participants who completed the pre- and post-program shared assessment in a community-centre, one assessor conducted the assessment in-person while another assessor simultaneously assessed the participant remotely. These conditions were chosen to reflect the different contexts in which falls prevention programs such as MVF are delivered. While some participants can access in-person assessments at a local community centre, others may rely entirely on remote assessments due to geographical location, mobility restrictions or limited availability to trained, French-speaking healthcare professionals. Since MVF is delivered both in-person and remotely, establishing the inter-rater reliability of both assessment and consistency between assessors is essential.

For the in-person assessments, the chair and space requirements were the same as the ones described above for the remote evaluations. The in-person assessor provided verbal instructions to each participant and demonstrated the test. Once this was well understood, the in-person assessor as well as the remote assessor simultaneously conducted the TUG and FTSTS, recording the time the participant required to complete each test. Each assessor recorded their obtained times independently, making sure not to reveal them to the other assessor to limit bias. Each test was performed twice. The TUG time was recorded after the second attempt only, while both attempts of the FTSTS were timed and recorded as per the standardized recommendations for these tests.

### Data analysis

We analyzed demographic data descriptively and used narrative analysis to report adverse events or technical difficulties encountered during remote assessments. We used intraclass correlation coefficients (ICCs) to assess inter-rater reliability for the TUG and FTSTS between two remote assessors (Koo & Li, 2016). For the FTSTS comparison, each assessor’s score was the mean of the two attempts they timed for each participant. We estimated inter-rater reliability between the in-person and remote assessors using a two-way model, placing importance on absolute agreement with a 95% confidence interval based on two raters per site using the average time recorded for two FTSTS attempts from each participant for inter-rater reliability (Koo & Li, 2016). For the TUG, we used a two-way model, placing importance on absolute agreement with a 95% confidence interval based on two raters per site for a single TUG score from each participant. The same process was used for the remote only data. Scores from each assessor were recorded independently and only revealed once assessments were completed to reduce potential inter-rater bias. As per the ICC reporting guidelines (Portney & Watkins, 2015), the ICC was interpreted as follows: values less than 0.5, between 0.5 and 0.75, between 0.75 and 0.9, and greater than 0.90 respectively being indicative of poor, moderate, good, and excellent reliability. We performed the statistical analyses using jamovi (version 2.6) [Computer Software].

Absolute agreement refers to the times recorded by each rater having to be identical for each participant (Koo & Li, 2016). Since both the TUG and FTSTS determine if a participant is at increased risk of falls based on a time threshold, raters’ absolute agreement is necessary, as timing differences could change whether a participant was deemed to have exceeded a specific cutoff time. Both raters should have consistently similar recorded times, identifying the same participants as being at increased risk of falls according to the tests’ time thresholds. Since the FTSTS was completed twice by participants, during each assessment, we used the mean of the two recorded times for analysis.

Concurrent agreement between raters for the remote TUG and FTSTS tests for each participant was analyzed by comparing results stemming from remote assessment to the in-person reference ratings. We used cut-offs of > 14 s for the TUG and >15 s for the FTSTS, as they have been shown for in-person assessments to be indicative thresholds for an increased risk of falls (Buatois et al., 2008; Shumway-Cook et al., 2000). These assessments thresholds are used as part of a larger established primary prevention program (Marche vers le futur)(O’Neil et al., 2024).

## Results

### Demographic characteristics

Sixty-seven participants from five provinces, Ontario (n=10), Prince Edward Island (n=3), Quebec (n=20), New Brunswick (n=30), and British Columbia (n=4), consented to the study and completed both pre- and post-program assessments. Twenty participants (30%) were from the province of Quebec and 47 participants (70%) were from francophone minority communities. Of the 67 participants, 61 completed the online demographics questionnaire, with 82% identifying as women, 95% indicating French as their native language and 96.7% indicating French as their preferred language. Five participants were between the ages of 50-59 (8%), 20 participants were between the ages of 60-69 (33%), 24 participants were between the ages of 70-79 (39%), 11 participants were between the ages of 80-89 (18%) and 1 participant was 90+ years of age (2%). Fifty-seven participants (85%) completed the assessments at a community centre with an in-person assessor and a remote assessor (MVF Salle), and 10 participants (15%) completed the assessments in their home. Of these 10 participants, six were evaluated simultaneously by two remote assessors. The remaining four were evaluated by one remote assessor because a second assessor was unavailable and were therefore excluded from the remote inter-rater reliability analysis. Overall, 80 participants were screened; four were excluded based on screening criteria, and nine dropped out of the program, without completing post program assessment.

### Overall adverse events

No adverse events linked to the type of assessment occurred during the sessions or were reported by assessors after the sessions for both the TUG or the FTSTS. No technological difficulties were experienced during data collection.

### Objective 1: Inter-rater reliability for remote only pre-and post-program assessments

Ten participants completed pre- and post-program functional mobility assessments from their homes with an assessor only present remotely, demonstrating feasibility and safety of conducting functional mobility assessments remotely with adults aged 55 years or older. Six out of these 10 participants (n=6) assessed at home were rated simultaneously by two remote assessors. TUG and FTSTS results for these participants are presented in Tables 1A and 1B, respectively. For the TUG, our results demonstrated overall excellent inter-rater reliability between two remote assessors pre-program and good post-program inter-rater reliability (Table 1A). For the FTSTS conducted by two remote assessors, our results demonstrated overall excellent inter-rater reliability between two remote assessors for both pre-program and post-program assessments (Table 1B). However, based on this small sample size, our findings should only be considered as preliminary results.

**Table 1.** Descriptive inter-rater reliability statistics of pre- and post-program remote assessment for the A. Timed Up and Go (TUG) and B. the Five Times Sit-to-Stand (FTSTS)

| **A. TUG** | **Preprogram Remote TUG (s) Assessor 1** | **Preprogram Remote TUG (s) Assessor 2** | **Post-program Remote TUG (s) Assessor 1** | **Post-program Remote TUG (s) Assessor 2** |
|---|---|---|---|---|
| **Mean time (±SD) (n=6)** | 10.80 (±2.05) | 11.10 (±1.90) | 10.80 (±2.30) | 10.40 (±1.74) |
| **Range (min-max) (n=6)** | 8.38-14.20 | 8.72-14.10 | 7.77-13.70 | 8.01-12.70 |
| **Inter-rater reliability (ICC) (n=6)** | 0.978 (CI 95% 0.892-1.12) |  | 0.833 (CI 95% 0.431-1.330) |  |
| **B. FTSTS** | **Preprogram Remote FTSTS (s) Assessor 1** | **Preprogram Remote FTSTS (s) Assessor 2** | **Post-program Remote FTSTS (s) Assessor 1** | **Post-program Remote FTSTS (s) Assessor 2** |
| **Mean time (±SD) (n=6)** | 12.60 (±2.79) | 13.30 (±2.15) | 10.80 (±1.22) | 11.20 (±1.58) |
| **Range (min-max) (n=6)** | 10.10-17.60 | 10.30-16.90 | 8.76-12.30 | 8.79-12.60 |
| **Inter-rater reliability (ICC) (n=6)** | 0.916 (CI 95% 0.375-1.53) |  | 0.937 (CI 95% 0.575-1.36) |  |

### Objective 2: Inter-rater reliability for in-person and remote pre-and post-program assessments

Fifty-seven participants completed pre- and post-program TUG and FTSTS at their local community centre, with one assessor conducting the assessment remotely and one assessor simultaneously conducting the assessment in-person. For the TUG, the preprogram inter-rater reliability between in-person and remote assessors quantifies using the single rating absolute agreement was 0.986 (95% CI 0.977-0.996) and 0.982 (95% CI 0.970-0.997) for the post-program agreement. This demonstrates overall excellent reliability between in-person assessment and remote assessment for both pre and post assessments for the TUG (Table 2A). For the FTSTS, the preprogram inter-rater reliability between in-person and remote assessors quantifies using the two-ways average absolute agreement was 0.997 (95% CI 0.989-1.000) and 0.987 (95% CI 0.954-0.998) for the post-program agreement. This demonstrates overall excellent reliability between in-person assessment and remote assessment for both pre and post assessments for the FTSTS (Table 2B).

**Table 2.** Descriptive and inter-rater reliability statistics of pre- and post-program assessment in-person versus remote for the A. TUG and the B. FTSTS.

| A. **TUG** | **Preprogram in-person TUG (s)** | **Preprogram Remote TUG (s)** | **Post-program in-person TUG (s)** | **Post-program Remote TUG (s)** |
|---|---|---|---|---|
| **Mean Time (±SD) (n=57)** | 8.81 (±1.98) | 8.78 (±1.93) | 8.53 (±1.88) | 8.50 (±1.87) |
| **Range (min-max) (n=57)** | 5.53-14.10 | 5.74-14.50 | 5.00-15.20 | 5.61-15.60 |
| **Inter-rater reliability (ICC)** | 0.986 (95% CI 0.977-0.996) |  | 0.982 (95% CI 0.970-0.997) |  |
| B. **FTSTS** | **Preprogram in-person FTSTS (s)** | **Preprogram Remote FTSTS (s)** | **Post-program in-person FTSTS (s)** | **Post-program Remote FTSTS (s)** |
| **Mean Time (±SD) (n=57)** | 11.60 (±2.76) | 11.50 (±2.67) | 10.50 (±2.35) | 10.30 (±2.23) |
| **Range (min-max) (n=57)** | 6.76-23.60 | 6.93-23.10 | 6.97 -18.50 | 7.00-17.80 |
| **Inter-rater reliability (ICC)** | 0.997 (95% CI 0.989-1.000) |  | 0.987 (95% CI 0.954-0.998) |  |

### Objective 3: Agreement between assessor for falls risk screening conducted remotely when compared to in-person assessments

Identification of individuals at increased risk of falls is based on interpretation of the FTSTS and TUG standardized tests thresholds. Out of the 57 participants assessed simultaneously in-person and remotely, a total of five out of 57 participants were identified as being at increased risk of falls preprogram, and two of those five participants remained at fall risk in post-assessment. For the TUG, one assessment out of 57 participants demonstrated agreement in scores above established cut-off scores allowing to identify an increased risk using both the in-person and the remote assessments during the pre- and post-program assessments. For the pre and post TUG assessment, Bland-Altman analysis demonstrates excellent agreement between in-person and remote methods. The 95% limit of agreement between methods is around ±0.7seconds indicates good consistency across samples, and a slight positive bias of +0.1 seconds suggest no changes in clinical meaning (figure 1).

**Figure 1.**
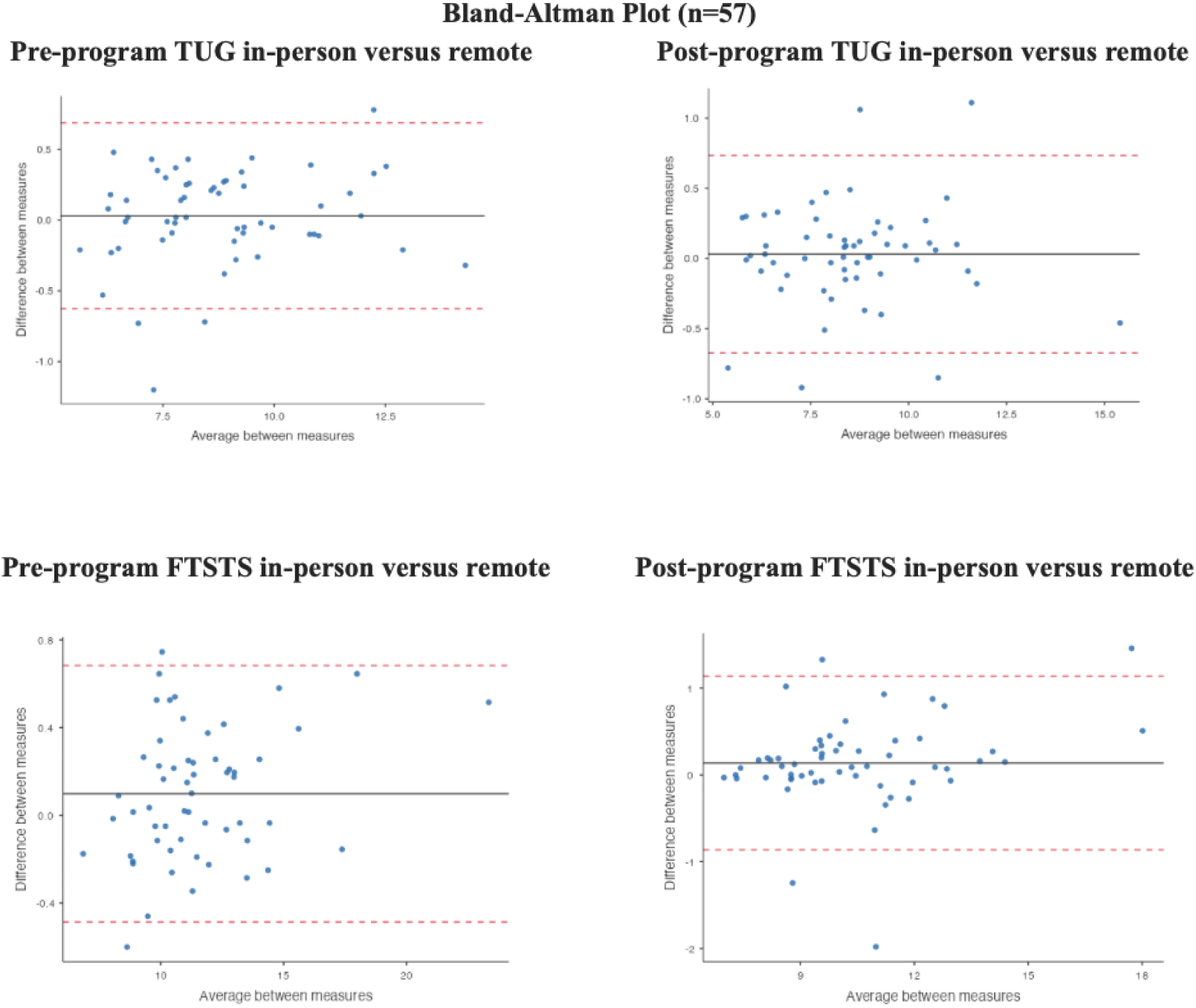
Bland-Altman plots of agreement between in-person and remote assessors’ scores for pre-program and post program TUG and FTSTS.

Similarly, for the FTSTS, simultaneous recorded times from the in-person preprogram assessment and remote assessment allowed to identify four out of 57 participants at increased risk of falls. Two out of 57 participants were identified at risk of falls during the post-program assessment using both in-person and remote assessments. In all instances, both the in-person and remote assessments identified the same individuals as being at increased risk of falls. For the FTSTS, Bland-Altman analysis demonstrates a slightly stronger agreement for the pre-assessment compared to the post-assessment methods with a slight positive bias of +0.1s suggesting no changes in clinical meaning. For the pre-assessment, most measurements were between a 95% limit of agreement of ±0.5s and 0.7 indicating good consistency across samples compared to an increased variability in post-assessment demonstrated by a 95% limit of agreement of ±0.9s and 1.1s (figure 1).

## Discussion

The main objectives of this study were to document the inter-rater reliability as well as the concurrent ratings agreement of balance and mobility screening for remote assessments and for remote compared with in-person assessments conducted in French-speaking communities. While assessing individuals presenting with functional mobility deficits in-person with a trained professional is the gold standard (Van Schooten et al., 2025), remote assessments may provide an option for individuals who face in-person access barriers (i.e., reduced mobility, transportation challenges, infectious diseases) (Anawade et al., 2024). Francophone communities have additional obstacles to healthcare access linked to language (Drolet et al., 2015; Mercure et al., 2018). Therefore, we found it was particularly relevant to examine the reliability and concurrent interrater agreement of remote functional mobility assessments in this specific underserved population.

Multiple studies have shown that a language barrier negatively impacts health outcomes (Belanger et al., 2023; De Moissac & Bowen, 2019; Drolet et al., 2015, 2017). Offering telehealth prevention programs in people’s preferred language allows individuals to access services they would otherwise be unable to receive and ensures that they are not excluded from innovation (O’Neil et al., 2024; Peng et al., 2024). Our study demonstrates that it is feasible to conduct remote dynamic balance and functional mobility assessments with French-speaking older adults, of whom 59% of participants were over 70 years of age, without compromising safety.

We found excellent inter-rater reliability, regardless of whether the TUG and FTSTS assessments were completed in-person or remotely. This suggests that for French-speaking communities, remote TUG and FTSTS assessments are a reliable alternative to in-person evaluations when the latter is not feasible. These findings therefore suggest that telerehabilitation may be a way to reduce inequities in access to healthcare amongst this population.

We considered concurrent agreement between remote and in-person assessors an important aspect to examine, not only to determine whether both assessors identified a similar number of participants as “at increased risk of falls,” but also whether they identified the same individuals. Our results show no disagreement in thresholds classification between the in-person and remote assessors’ recorded times. In total, five individuals were identified as being at risk of falls, having exceeded the in-person reference rating time thresholds of 14 s for the TUG and 15 s for the average of two FTSTS. Recorded times from both assessors were consistent, with in-person and remote assessments identifying the same five individuals. Our findings align with concurrent agreement results from a recent study by van Schooten et al. (2025), suggesting that remote assessment is similar to that of in-person TUG and FTSTS which supports its use for fall risk screening (Bohannon et al., 2015; Buatois et al., 2008; Shumway-Cook et al., 2000; Van Schooten et al., 2025). This information is critical for the implementation of telehealth programs since it provides strong evidence that French-speaking older adults living in remote areas can effectively participate in valid and reliable assessments to confirm or refute eligibility for fall-prevention programs to which they would otherwise not have access.

Beyond fall-prevention, our findings contribute to the larger context of ageism and digital health. There is an increased demand on healthcare systems due to an increase in individuals experiencing chronic health conditions and comorbidities associated with aging, leading to mobility limitations (Chen et al., 2023; Jane Osareme, Ogugua et al., 2024). Digital health may provide an effective option to help alleviate healthcare demands by increasing access and efficiency of care (Willems et al., 2021). However, beliefs rooted in ageism are still common (Mace et al., 2022). Older adults are often perceived as having more difficulties with the adequate use of digital health (Mace et al., 2022). This has caused older adults to be disproportionately disadvantaged or excluded from digital health service options compared to younger individuals, causing a “digital divide” and healthcare inequities (Mace et al., 2022). Our findings suggest safety, reliability, and validity of remote falls risk assessments in French-speaking older adults, supporting the idea that older adults can actively engage and participate in remote assessments and should have the option to do so (Peng et al., 2024). Supported by the Mohsin et al. (2025) taxonomy (Mohsin et al., 2025), future directions in this area of remote assessment could be used for individuals living with neurological conditions.

### Strengths and limitations

A notable strength of this study is the significant sample size, representing multiple geographical regions (5 provinces) and various assessment settings (home-based, local community centres). Since French-speaking older adults are often underserved in their language of choice, assessing the concurrent ratings agreement and reliability of assessments while considering linguistic factors is important, as linguistic barriers can negatively affect outcomes (De Moissac & Bowen, 2019). All remote TUG and FTSTS tests were assessed by physiotherapists who are experienced in telerehabilitation. Further research on inter-rater reliability conducted by healthcare workers who are not as familiar with remote assessments would be interesting. Self-reported, anonymous data collection regarding participant demographics also constituted a limitation. Each participant was instructed to fill out an online demographic questionnaire via SurveyMonkey, however, since this questionnaire was anonymous, it was not possible for us to identify which participants had not completed it. An alternate way of collecting this information would prevent future loss of useful research data.

Because this study was pan-Canadian, it was not possible to have initial practice assessment sessions with two assessors present in-person to establish a baseline prior to beginning data collection. We based the in-person inter-rater validity and reliability on previous studies that had already established excellent values between raters for this assessment (Buatois et al., 2008; Christopher et al., 2021; Whitney et al., 2005). In order to optimize standardization before the start of data collection, assessors watched TUG and FTSTS practice videos and discussed procedures such as specific instructions and prompts to be given to participants, when to begin and stop timers for the different tests, ideal camera placement and other required equipment to ensure safety and make sure participants would always be seen from head to toe throughout the assessments.

While 57 participants completed assessments comparing in-person and simultaneous remote evaluations, only 10 participants underwent entirely remote assessments because in-person evaluations were not available in their region. Of these 10 participants, four were evaluated by only one assessor due to the unavailability of a second assessor on the day of assessment. Therefore, only 6 participants were assessed simultaneously by two remote assessors, allowing inter-rater reliability and concurrent interrater agreement to be examined in the fully remote condition. Although our findings demonstrated excellent inter-rater reliability and concurrent interrater agreement, we consider our results preliminary as a larger sample size for fully remote assessments would strengthen these conclusions.

Finally, Internet was accessible and high-speed in all regions where the remote assessments were completed. Areas that do not have access to such consistent Internet services would likely pose challenges to the feasibility and reliability of the tests conducted remotely.

### Clinical considerations

Results from this study showcased reliability of conducting an assessment simultaneously by two assessors, one in-person, and one remote. However, other types of reliability, for example remote-to-remote repeatability across days, would have to be confirmed. While remote assessments are a valid and reliable option to improve access to falls risk screening, it is imperative that client safety remains a priority. Whether the service is delivered in-person or remotely, clinical reasoning as well as person-centered care should remain central. Our study suggests the following recommendations to reduce safety risks while conducting remote assessments: 1) sending instructions to the participant prior to the assessment detailing the equipment, space and screen placement requirements; 2) having another person physically present with the participant when possible; 3) educating participants on how to get up from a fall; 4) properly documenting emergency contact information prior to conducting a remote screening; 5) training healthcare providers to conduct safe remote assessments, emphasizing the importance of giving clear instructions to complete tasks near a wall, chair or other solid object the participant can hold onto in case of a loss of balance. While this provides useful feasibility information, future studies including people with reduced mobility and higher risk of falls are necessary to further investigate safety within remote assessment context.

## Conclusion

Recent research has shown that remote balance and mobility assessments have good reliability and validity. However, these findings had not yet been explored with French-speaking older adults. Our study adds to the literature by demonstrating excellent agreement between simultaneous remote assessments and in-person community-led TUG and FTSTS assessments. Our results suggest that simultaneous remote assessments of the TUG and FTSTS may be a reliable option to increase availability of health care services in a language of choice for Francophone older adults in Canada. Recognizing language preferences and ensuring inclusion of older adults in digital health innovations, such as remote assessments is critical to reduce health inequities.

## Data Availability

All data produced in the present study are available upon reasonable request to the authors

## References

Anawade, P. A., Sharma, D., & Gahane, S. (2024). A Comprehensive Review on Exploring the Impact of Telemedicine on Healthcare Accessibility. Cureus, 16(3), e55996. 10.7759/cureus.55996

Belanger, C., Carr, K., Peixoto, C., & Bjerre, L. M. (2023). Distance, access and equity: A cross-sectional geospatial analysis of disparities in access to primary care for French-only speakers in Ottawa, Ontario. CMAJ Open, 11(3), E434–E442. 10.9778/cmajo.20220061

Bohannon, R. W., Wang, Y.-C., & Gershon, R. C. (2015). Two-Minute Walk Test Performance by Adults 18 to 85 Years: Normative Values, Reliability, and Responsiveness. Archives of Physical Medicine and Rehabilitation, 96(3), 472–477. 10.1016/j.apmr.2014.10.006

Buatois, S., Miljkovic, D., Manckoundia, P., Gueguen, R., Miget, P., Vançon, G., Perrin, P., & Benetos, A. (2008). Five times sit to stand test is a predictor of recurrent falls in healthy community-living subjects aged 65 and older. Journal of the American Geriatrics Society, 56(8), 1575–1577. 10.1111/j.1532-5415.2008.01777.x

Chen, C., Ding, S., & Wang, J. (2023). Digital health for aging populations. Nature Medicine, 29(7), 1623–1630. 10.1038/s41591-023-02391-8

Christopher, A., Kraft, E., Olenick, H., Kiesling, R., & Doty, A. (2021). The reliability and validity of the Timed Up and Go as a clinical tool in individuals with and without disabilities across a lifespan: A systematic review: Psychometric properties of the Timed Up and Go. Disability and Rehabilitation, 43(13), 1799–1813. 10.1080/09638288.2019.1682066

De Moissac, D., & Bowen, S. (2019). Impact of Language Barriers on Quality of Care and Patient Safety for Official Language Minority Francophones in Canada. Journal of Patient Experience, 6(1), 24–32. 10.1177/2374373518769008

Drolet, M., Arcand, I., Benoît, J., Savard, J., Savard, S., & Lagacé, J. (2015). Agir pour avoir accès à des services sociaux et de santé en français: Des Francophones en situation minoritaire nous enseignent quoi faire! Canadian Social Work Review, 32(1–2), 5–26. 10.7202/1034141ar

Drolet, M., Bouchard, P., & Savard, J. (Eds.). (2017). Accessibilité et offre active: Santé et services sociaux en contexte linguistique minoritaire. University of Ottawa Press. 10.2307/j.ctv5vdcp0

Farzad, M., MacDermid, J., Ferreira, L., Szekeres, M., Cuypers, S., & Shafiee, E. (2023). A description of the barriers, facilitators, and experiences of hand therapists in providing remote (tele) rehabilitation: An interpretive description approach. Journal of Hand Therapy, *3c*(4), 805–816. 10.1016/j.jht.2023.06.004

Fauchard, T., & Le Cren, F. (2009). Présentation du programme intégré d’équilibre dynamique (PIED). Science & Sports, 24(3–4), 152–159. 10.1016/j.scispo.2007.01.008

Franco, J. B., Maximino, L. P., Secchi, L. L. B., Antonelli, B. C., & Blasca, W. Q. (2024). What Are the Barriers to Telerehabilitation in the Treatment of Musculoskeletal Diseases? Portuguese Journal of Public Health, 42(1), 33–42. 10.1159/000534762

González-Calvo, J., González, V. M., & Lorig, K. (1997). Cultural diversity issues in the development of valid and reliable measures of health status. Arthritis Care and Research: The Official Journal of the Arthritis Health Professions Association, 10(6), 448–456. 10.1002/art.1790100613

International Test Commission (ITC). (2019). ITC Guidelines for the Large-Scale Assessment of Linguistically and Culturally Diverse Populations. International Journal of Testing, 19(4), 301–336. 10.1080/15305058.2019.1631024

Jacobson, C. L., Foster, L. C., Arul, H., Rees, A., & Stafford, R. S. (2021). A Digital Health Fall Prevention Program for Older Adults: Feasibility Study. JMIR Formative Research, 5(12), e30558. 10.2196/30558

Jane Osareme, Ogugua, Muridzo Muonde, Chinedu Paschal Maduka, Tolulope O Olorunsogo, & Olufunke Omotayo. (2024). Demographic shifts and healthcare: A review of aging populations and systemic challenges. International Journal of Science and Research Archive, 11(1), 383–395. 10.30574/ijsra.2024.11.1.0067

Jasper, A., Karim, R., Vitente, A. C., Rafael, C. (Minnie), Tayag, E., Uy, S. J. M., Baloy, R. K., & Lazaro, R. (2025). Concurrent Validity and Reliability of In-Person and Supervised Remote STEADI Fall Risk Assessment in Community-Dwelling Older Adults. Journal of Geriatric Physical Therapy, 48(3), 116–124. 10.1519/JPT.0000000000000446

Koo, T. K., & Li, M. Y. (2016). A Guideline of Selecting and Reporting Intraclass Correlation Coefficients for Reliability Research. Journal of Chiropractic Medicine, 15(2), 155– 163. 10.1016/j.jcm.2016.02.012

Lee, A. C., Deutsch, J. E., Holdsworth, L., Kaplan, S. L., Kosakowski, H., Latz, R., McNeary, L. L., O’Neil, J., Ronzio, O., Sanders, K., Sigmund-Gaines, M., Wiley, M., & Russell, T. (2024). Telerehabilitation in Physical Therapist Practice: A Clinical Practice Guideline From the American Physical Therapy Association. Physical Therapy, 104(5), pzae045. 10.1093/ptj/pzae045

Mace, R. A., Mattos, M. K., & Vranceanu, A.-M. (2022). Older adults can use technology: Why healthcare professionals must overcome ageism in digital health. Translational Behavioral Medicine, 12(12), 1102–1105. 10.1093/tbm/ibac070

Mercure, D., Côté, I., & Garceau, M.-L. (2018). Santé et accès aux soins de santé des communautés francophones en situation minoritaire au Canada (CFSM). Reflets : revue d’intervention sociale et communautaire, 24(2), 10–14. 10.7202/1053861ar

Mohammed, H. T., Hyseni, L., Bui, V., Gerritsen, B., Fuller, K., Sung, J., & Alarakhia, M. (2021). Exploring the use and challenges of implementing virtual visits during COVID-19 in primary care and lessons for sustained use. PLOS ONE, 16(6), e0253665. 10.1371/journal.pone.0253665

Mohsin, S. S., Salman, O. H., Jasim, A. A., Al-Nouman, M. A., & Kairaldeen, A. R. (2025). A systematic review on the roles of remote diagnosis in telemedicine system: Coherent taxonomy, insights, recommendations, and open research directions for intelligent healthcare solutions. Artificial Intelligence in Medicine, 160, 103057. 10.1016/j.artmed.2024.103057

Montero-Odasso, M., Van Der Velde, N., Martin, F. C., Petrovic, M., Tan, M. P., Ryg, J., Aguilar-Navarro, S., Alexander, N. B., Becker, C., Blain, H., Bourke, R., Cameron, I. D., Camicioli, R., Clemson, L., Close, J., Delbaere, K., Duan, L., Duque, G., Dyer, S. M., … Rixt Zijlstra, G. A. (2022). World guidelines for falls prevention and management for older adults: A global initiative. Age and Ageing, 51(9), afac205. 10.1093/ageing/afac205

Moriichi, K., Fujiya, M., Ro, T., Ota, T., Nishimiya, H., Kodama, M., Yoshida, N., Hattori, Y., Hosokawa, T., Hishiyama, H., Kunimoto, M., Hayashi, H., Hirokawa, H., & Yoshida, A. (2022). A novel telerehabilitation with an educational program for caregivers using telelecture is feasible for fall prevention in elderly people: A case series. Medicine, 101(6), e27451. 10.1097/MD.0000000000027451

O’Neil, J., Dionne, N., Marchand, S., Cardinal, D., Handrigan, G., & Savard, J. (2024). Reach, Adoption, and Implementation Strategies of a Telehealth Fall Prevention Program: Perspectives From Francophone Communities Across Canada. Health Promotion Practice, 15248399241252807. 10.1177/15248399241252807

Patterson, P. B., Roddick, J., Pollack, C. A., & Dutton, D. J. (2022). Virtual care and the influence of a pandemic: Necessary policy shifts to drive digital innovation in healthcare. Healthcare Management Forum, 35(5), 272–278. 10.1177/08404704221110084

Pelicioni, P. H. S., Waters, D. L., Still, A., & Hale, L. (2022). A pilot investigation of reliability and validity of balance and gait assessments using telehealth with healthy older adults. Experimental Gerontology, 162, 111747. 10.1016/j.exger.2022.111747

Peng, W., Zhu, G., Chen, Z., Hou, T., Luo, Y., Huang, L., Qiao, J., & Li, Y. (2024). Digital Technology Use in US Community-Dwelling Seniors With and Without Homebound Status. Journal of the American Medical Directors Association, 25(11), 105284. 10.1016/j.jamda.2024.105284

Pillay, J., Gaudet, L. A., Saba, S., Vandermeer, B., Ashiq, A. R., Wingert, A., & Hartling, L. (2024). Falls prevention interventions for community-dwelling older adults: Systematic review and meta-analysis of benefits, harms, and patient values and preferences. Systematic Reviews, 13(1), 289. 10.1186/s13643-024-02681-3

Portney, L. G., & Watkins, M. P. (2015). Foundations of clinical research: Applications to practice (3rd edition). F.A. Davis Company.

Savard, J., Labossière, S., Cardinal, D., Pinet, B., & Borris, C. (2018). Évaluation de Marche vers le futur, un programme novateur de prévention des chutes offert par videoconference. Canadian Journal on Aging / La Revue Canadienne Du Vieillissement, 37(4), 363–376. 10.1017/S0714980818000326

Scott, V., Gallagher, E., Higginson, A., Metcalfe, S., & Rajabali, F. (2011). Evaluation of an evidence-based education program for health professionals: The Canadian Falls Prevention Curriculum© (CFPC). Journal of Safety Research, 42(6), 501–507. 10.1016/j.jsr.2011.10.004

Shumway-Cook, A., Brauer, S., & Woollacott, M. (2000). Predicting the probability for falls in community-dwelling older adults using the Timed Up & Go Test. Physical Therapy, 80(9), 896–903.

Taing, D., & McKay, K. (2017). Better Strength, Better Balance! Partnering to deliver a fall prevention program for older adults. Canadian Journal of Public Health, 108(3), e314–e319. 10.17269/CJPH.108.5901

Van Schooten, K. S., Steffens, D., Engeler, A., Letton, M., Brown, A., Perram, A., Miles, L., Ngo, M., & Delbaere, K. (2025). Remote assessment of physical function in older people: Feasibility, safety and agreement with in-person administration. Age and Ageing, 54(9), afaf266. 10.1093/ageing/afaf266

Whitney, S. L., Wrisley, D. M., Marchetti, G. F., Gee, M. A., Redfern, M. S., & Furman, J. M. (2005). Clinical measurement of sit-to-stand performance in people with balance disorders: Validity of data for the Five-Times-Sit-to-Stand Test. Physical Therapy, 85(10), 1034–1045.

Willems, S. H., Rao, J., Bhambere, S., Patel, D., Biggins, Y., & Guite, J. W. (2021). Digital Solutions to Alleviate the Burden on Health Systems During a Public Health Care Crisis: COVID-19 as an Opportunity. JMIR mHealth and uHealth, 9(6), e25021. 10.2196/25021

